# Transcriptomic analyses of human brains with Alzheimer’s disease identified dysregulated epilepsy-causing genes

**DOI:** 10.1101/2025.01.02.25319900

**Authors:** Abdallah M. Eteleeb, Suélen Santos Alves, Stephanie Buss, Mouhsin Shafi, Daniel Press, Norberto Garcia-Cairasco, Bruno A. Benitez

## Abstract

**Background & Objective:** Alzheimer’s Disease (AD) patients at multiple stages of disease progression have a high prevalence of seizures. However, whether AD and epilepsy share pathophysiological changes remains poorly defined. In this study, we leveraged high-throughput transcriptomic data from sporadic AD cases at different stages of cognitive impairment across multiple independent cohorts and brain regions to examine the role of epilepsy-causing genes.

**Methods:** Epilepsy-causing genes were manually curated, and their expression levels were analyzed across bulk transcriptomic data from three AD cohorts and three brain regions. RNA-seq data from sporadic AD and control cases from the Knight ADRC, MSBB, and ROSMAP cohorts were processed and analyzed under the same analytical pipeline. An integrative clustering approach employing machine learning and multi-omics data was employed to identify molecularly defined profiles with different cognitive scores.

**Results:** We found several epilepsy-associated genes/pathways significantly dysregulated in a group of AD patients with more severe cognitive impairment. We observed 15 genes consistently downregulated across the three cohorts, including sodium and potassium channels, suggesting that these genes play fundamental roles in cognitive function or AD progression. Notably, we found 25 of these genes dysregulated in earlier stages of AD and become worse with AD progression.

**Conclusion:** Our findings showed that epilepsy-causing genes showed changes in the early and late stages of AD progression, suggesting that they might be playing a role in AD progression. We can not establish directionality or cause-effect with our findings. However, changes in the epilepsy-causing genes might underlie the presence of seizures in AD patients, which might be present before or concurrently with the initial stages of AD.

## Introduction

Evidence from studies involving rodents and humans across various research methods strongly supports the idea that network dysconnectivity and hyperexcitability are important intrinsic features of Alzheimer’s disease (AD) [1]. Seizures and epileptiform activity are classic manifestations of network hyperexcitability [2]. AD is associated with an increased risk of seizures, with up to 15–20% of patients affected. Seizures are associated not only with an earlier onset of cognitive decline but also with more rapid disease progression [2]. Specific features of epileptiform discharges are associated with clinical seizures in AD. Epileptiform activity and chronic hyperexcitability promote amyloid-β (Aβ) plaque deposition and tau hyperphosphorylation [3].

A recent meta-analysis identified a 1.8-fold increased risk of AD in people with epilepsy, while patients with AD are at a 3.1-fold higher risk of epilepsy [4]. Recent studies have shown an important bidirectional connection between epilepsy and AD [5–9] and highlighted that cognitive impairment frequently coexists with epilepsy and appears to be associated with multiple phenotype variables including age, gender, educational attainment, and the dosage of antiseizure medications [10]. The rate of dementia incidence among epilepsy patients has been shown to increase, particularly among the elderly [5,11]. This relationship has been further reported to be associated with worse clinical outcomes and shorter lifespans [12,13]. Furthermore, increasing evidence suggests that people who develop epilepsy after the age of 40, without a known cause, are more likely to develop AD compared to those who do not have epilepsy [5]. The association between AD and epilepsy, particularly mesial temporal lobe epilepsy, has led researchers to identify a potential new subtype of AD characterized by more substantial neuronal loss and a rapid decline in cognitive and clinical functions [8]. A causal association between AD and generalized epilepsy was observed, and this association was robust to sensitivity analyses. It was further validated by a significant association between lower CSF Aβ42 and an increased risk of generalized epilepsy [6].

Previous studies have investigated the correlation between AD pathology and epilepsy [3,14–27] and have identified the underlying factors that have implications in this correlation. Aβ plaques and neurofibrillary tangles (NFTs), the two major hallmarks of AD, have been frequently linked to epilepsy [3,14,15,17]. Additionally, APOE ℇ4, the most significant genetic risk factor for sporadic AD, was also identified as a contributing factor for epilepsy [28]. Brain proteomics from epilepsy and AD patients have identified alterations in numerous proteins that interact with tau or are regulated by tau expression in epilepsy [20] although the exact mechanisms remain unclear. Other common factors between epilepsy and AD are changes in the glutamate and glutamine cycle [21,22], structural or functional defects of ion channel proteins [23], neuroinflammation [24], locus coeruleus degeneration [25] and metabolic alterations [26,27]. However, the molecular changes underlying the link between AD and epilepsy remain poorly understood. In this study, we analyzed high-throughput transcriptomic profiles from multiple AD cohorts and brain regions to investigate whether shared transcriptomic signatures can provide novel insights into the role epilepsy-causing genes in AD.

## Materials and Methods

### Study cohorts

#### The Knight Alzheimer Disease Research Center (Knight ADRC)

Summary statistics from the transcriptomic data from the parietal cortex from 255 sporadic AD participants and 23 controls were included in this study [29]. Total RNA was extracted from frozen parietal cortex tissue using a Tissue Lyser LT and purified using RNeasy Mini Kits (Qiagen). The Nanodrop 8000 (Thermo Scientific) and TapeStation 4200 (Agilent Technologies) were used to perform quality control of the RNA’s concentration, purity, and degradation. The RNA integrity number (RIN) was calculated using an RNA 6000 Pico assay on a Bioanalyzer 2100 and TapeStation 4200 (Agilent Technologies). The DV200 value is defined as the percentage of nucleotides greater than 200nt. All cDNA libraries were prepared using a TruSeq Stranded Total RNA Sample Prep with Ribo-Zero Gold kit (Illumina) and sequenced on an Illumina HiSeq 4000 using 2 × 151 paired-end reads at the McDonnell Genome Institute at Washington University in St. Louis.

#### The Mount Sinai Brain Bank (MSBB) study

The RNA-seq raw data from the MSBB study [30] was downloaded from the Synapse portal (syn3157743). Only data from the parahippocampal gyrus (PHG, BA36) were selected for this study. Briefly, RNA-seq libraries were prepared using the TruSeq RNA Sample Preparation Kit v2 (Illumina, San Diego, California, USA). The rRNAs were depleted using the Ribo-Zero rRNA Removal Kit (human/mouse/rat) (Illumina). Single-end non-standard reads of 101 bp were generated by Illumina HiSeq 2500 (Illumina).

#### The Religious Orders Study and Memory and Aging Project (ROSMAP)

The RNA-seq raw data from the dorsolateral prefrontal cortex (DLPFC) from the ROSMAP Study [16] were downloaded from the Synapse portal (syn17008934). Briefly, RNA was extracted using Qiagen’s miRNeasy mini kit (cat. no. 217004) and the RNase-free DNase Set (cat. no. 79254) and quantified by Nanodrop. Agilent Bioanalyzer evaluated quality. Library preparation was performed by poly-A selection followed by first strand-specific cDNA synthesis, then dUTP for second strand-specific cDNA synthesis, and fragmentation and Illumina adapter ligation for library construction. Illumina HiSeq generated paired-end read sequences with a length of 101 bp. Only No Cognitive Impairment (NCI) and AD with no other cause of cognitive impairment cases were used in this study [29].

### Statistical Analysis

#### RNA-seq QC, alignment, and gene expression quantification

RNA-seq datasets from the three cohorts were processed and aligned using our in-house RNA-seq pipeline [29]. Reference genome GRCh38 and GENCODE 33 annotation, including the addition of ERCC spike-in annotations, were used. Before alignment, the quality of raw read sequences for all libraries was assessed using FastQC (v0.11.9) [31]. All read sequences were aligned to the human reference genome (GRCh38) using STAR (v.2.7.1a) [32]. Post-alignment quality was evaluated using Picard tools (v.2.8.2) [33]. All samples that failed to pass the QC or were outliers were removed from the downstream analyses. Raw read counts for transcripts/genes were generated using STAR, and normalized gene expression levels were computed in Fragments Per Kilobase of transcript per Million mapped reads (FPKM) format as previously described [29].

#### Molecular profiling using an integrative clustering approach

AD molecular profiles were identified using iClusterBayes [34], iClusterPlus R package (v1.22.0), [35]. Differential expression (DE) analyses were performed to compare cases in each AD profile to the control cases and the cases in other profiles using DESeq2 R package (v.1.22.2) [36]. All models were adjusted by sex, age at death, and the percentage of astrocytes and neurons. Only genes with expression > 0.5 CPM (count per million) in at least 25% of samples in either group being compared were retained for downstream analyses. All genes with FDR < 0.05 were considered differentially expressed genes (DEGs) [29]. This study only focuses on the DEGs between the worse cognitive profile and other sporadic AD cases in each cohort.

#### Pathways enrichment Analysis

Pathway analyses were performed using the EnrichR R package (v3.0) [180], using dysregulated genes to compare the worse cognitive profile of the sporadic AD cases across the three cohorts.

#### Brain Cell proportion based on single-cell transcriptomics

The estimated cellular population structure from gene expression data was computed using the CellMix R package (v1.6.2) [37]. The gene panel (marker genes expressed highly in specific cell types) and the machine learning model[38].

## Results

### Dysregulated epilepsy-causing genes in the multi-omics profile associated with synaptic plasticity dysfunction in AD

We have recently identified a subtype of AD cluster associated with significant dysregulation of synapse-related genes and pathways (from now on referred to as “*synaptic plasticity dysfunction or SPD*”). This subtype has clinical and neuropathological features correlates including significantly higher clinical dementia rating (CDR) at death, shorter survival after symptom onset, more severe neurodegeneration and astrogliosis, and decreased levels of metabolomic, suggesting an association with more severe cognitive profiles [29]. Here, we extended these analyses to study the correlation between the SPD profile and epilepsy-causing genes using transcriptomic profiles from AD patients (**Table 1**). We combined a list of 105 genes shown by the Epilepsy Foundation (https://www.epilepsy.com/causes/genetic) and Wang et al., 2017 [39]. We integrated it with the multi-omics integration analyses we performed previously [29].

**Table 1.** Demographic characteristics of AD and control cases.

| Study | N | Tissue<sup>a</sup> | AD |  |  | Control |  |  |
| --- | --- | --- | --- | --- | --- | --- | --- | --- |
|  |  |  | Female (%) | Age (mean,sd) | APOE ε4+ (%) | Female (%) | Age (mean,sd) | APOE ε4+ (%) |
| Knight ADRC | 278 | PC | 60 | 83.8(8.6) | 58.04 | 73.9 | 88.6(8.8) | 0 |
| MSBB | 217 | BM36 | 67.1 | 84.4(6.7) | 19.29 | 66.6 | 80.6(8.3) | 18.5 |
|  |  | BM10 | 67 | 84.2(6.8) | 20.73 | 69.7 | 81.3(7.0) | 15.1 |
|  |  | BM22 | 62.9 | 83.6(6.9) | 23.38 | 70 | 81(8.0) | 16.6 |
|  |  | BM44 | 66.6 | 84.3(6.7) | 20.92 | 64.2 | 80.5(8.3) | 17.8 |
| ROSMAP | 237 | DLPFC | 70.8 | 90.7(5.9) | 39.58 | 53.7 | 84.1(6.8) | 10.7 |

In our previous work [29], we performed multiple differential gene expression analyses (see Material and Methods) to compare cases in the SPD to other AD cases (from now on referred to as “sporadic AD or sAD”, **Fig. 1a-c**) as well as to the control cases in each AD cohort. We focused our analyses on the epilepsy-causing genes dysregulated between the SPD and the sAD profile. We first intersected the list of these genes with the list of dysregulated genes between the SPD and sAD profile in the three cohorts (**Fig. 1d**). Out of the 105 epilepsy-associated genes we selected, 45 (43%) were significantly dysregulated in the Knight ADRC SPD, 67 (64%) in the MSBB profile, and 41 (39%) in the ROSMAP profile (**Fig. 1d**). We then extracted the shared set of dysregulated genes across the three cohorts which included 16 epilepsy-associated genes (**Fig. 1d** and **Table 2**). Interestingly, 15 genes were consistently downregulated across the three cohorts, and only one gene (*CERS1*) was upregulated (**Fig. 1e-g**). Using RNA-seq data from human induced pluripotent stem cells (iPSC)-derived neurons, microglia, macrophages, and astrocytes (see Material and Methods), we observed that many of these genes were highly expressed in iPSC-derived neurons (**Fig. 2a**). These results were consistent with human brain gene expression generated from single-nuclei RNA-seq data from the parietal cortex of 67 Knight ADRC participants [40] (**Fig 2b**). Nine of these genes (*ARHGEF9, GABRA1, GABRB3, GABRG2, GRIN2A, HCN1, NECAP1, SCN1A, SCN8A*) were mainly synaptic genes. **Table 2** shows the different epileptic phenotypes in each of the 16 genes involved. For instance, *CERS1* was shown to be involved in progressive myoclonic epilepsies (PME) [41,42], a group of heterogeneous disorders characterized by myoclonus, epileptic seizures, and progressive neurologic deterioration, which may include cognitive decline and ataxia [43]. This may indicate the correlation between the cognitive profile of AD and epilepsy since people with PME show a decline in cognitive function over time. We observed that nine genes: *UBA5, ARHGEF9, GABRB3, SCN8A, HCN1, SLC25A12, NECAP1, FGF12, GABRA1*, were involved in the Early infantile epileptic encephalopathy (EIEE) phenotype. These results suggest that the SPD of AD involved multiple epilepsy-causing genes with multiple epilepsy phenotypes.

**Fig. 1:**
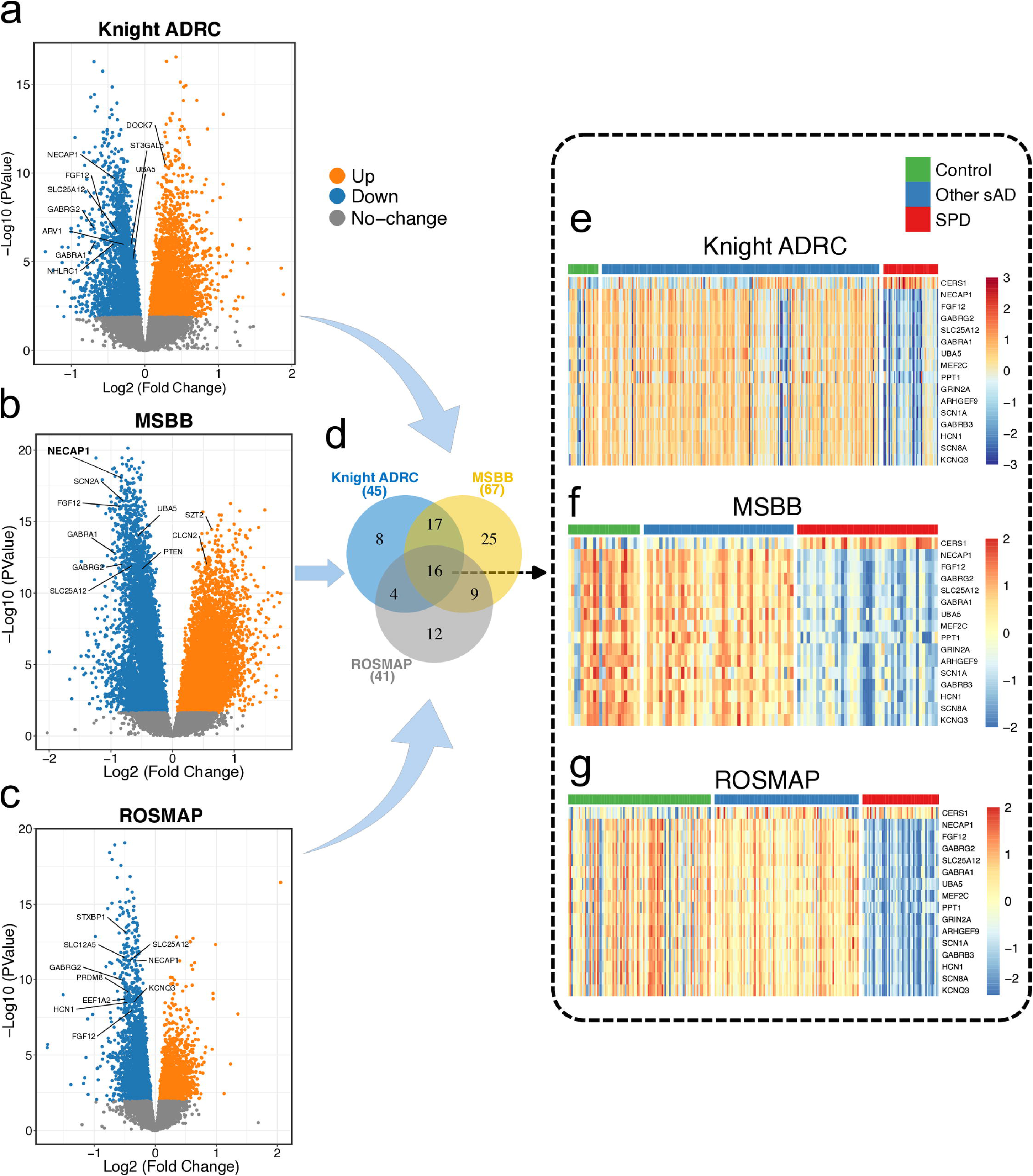
Dysregulated epilepsy-causing genes in the multi-omics profile associated with synaptic plasticity dysfunction (SPD) in AD. (**a**) Volcano plot showing the up-and downregulated genes identified between cases in the SPD profile vs other sporadic AD cases in the Knight ASRC cohort. (**b** and **c**) Same as “a” but in the MSBB and ROSMAP cohorts. (**c**) same as “a” and “b” but for ROSMAP cohort. (**d**) Venn diagrams depicting the number of dysregulated epilepsy-causing genes in each cohort as well as the overlap between them in three cohorts. (**e**) Heatmap of the expression profiles of the common epilepsy-causing genes from the knight ADRC cohort. (**f** and **g**) Same for “**e**” but from the MSBB and ROSMAP cohorts.

**Fig. 2:**
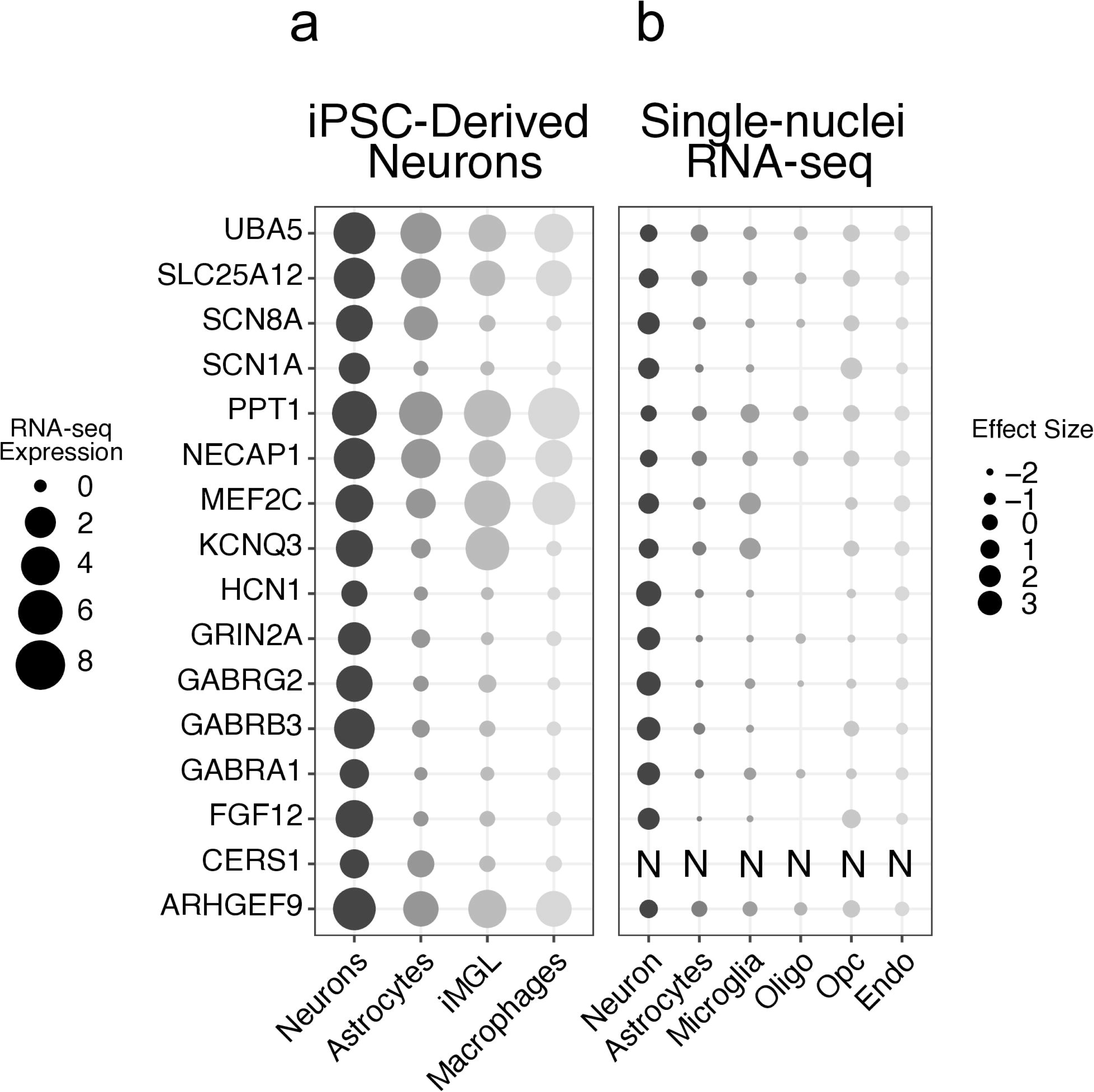
Dysregulated epilepsy-causing genes in the SPD profile are highly expressed in neurons. (**a**) The RNA-seq expression of the dysregulated epilepsy-causing genes in human induced pluripotent stem cells (iPSC)-derived neurons. (**b**) The effect size of the overepxression of each of the dysregulated epilepsy-causing gene in a speicfic cell type relative to other cell types computed from single-nuclei data from the Knight ADRC participants. “N” indicates that no data was available for CERS1 in the single-nuclei data.

**Table 2.** Summary of the statistical information for the shared epilepsy genes.

| Gene | Knight ADRC |  | MSBB |  | ROSMAP |  | Directionality of change | Phenotype |
| --- | --- | --- | --- | --- | --- | --- | --- | --- |
|  | FC* | FDR | FC | FDR | FC | FDR |  |  |
| CERS1 | 0.27 | 1.1e-03 | 0.67 | 3.7e-06 | 0.18 | 4.8e-02 | Up | Progressive myoclonic epilepsies (PME) [39,41,42] |
| UBA5 | -0.16 | 1.6e-04 | -0.56 | 1.1e-12 | -0.14 | 4.7e-02 | Down | Early infantile epileptic encephalopathy (EIEE) [140–142] |
| KCNQ3 | -0.23 | 4.8e-02 | -0.38 | 5.5e-06 | -0.35 | 3.0e-07 | Down | Benign familial neonatal seizures (BFNS) [101,143,144] |
| PPT1 | -0.24 | 4.9e-04 | -0.41 | 3.5e-08 | -0.11 | 3.7e-02 | Down | Neuronal ceroid lipofuscinosis [109,111,145] |
| ARHGEF9 | -0.25 | 9.4e-04 | -0.34 | 1.5e-05 | -0.18 | 4.2e-04 | Down | EIEE [146] |
| GABRB3 | -0.25 | 2.5e-02 | -0.43 | 1.5e-06 | -0.28 | 1.7e-05 | Down | Childhood absence epilepsy (CAE) [147,148]/EIEE [149] |
| SCN8A | -0.25 | 4.1e-02 | -0.3 | 2.6e-04 | -0.23 | 8.6e-04 | Down | Benign familial infantile seizures (BFIS) [150]/EIEE [151] |
| HCN1 | -0.3 | 3.2e-02 | -0.75 | 1.0e-10 | -0.46 | 3.2e-07 | Down | EIEE [65] |
| SLC25A12 | -0.37 | 8.6e-06 | -0.66 | 6.2e-11 | -0.42 | 1.4e-09 | Down | EIEE [152,153] |
| NECAP1 | -0.4 | 4.3e-08 | -0.83 | 1.0e-15 | -0.34 | 1.4e-09 | Down | EIEE [89] |
| SCN1A | -0.4 | 2.9e-03 | -0.62 | 2.5e-05 | -0.34 | 3.3e-05 | Down | Dravet syndrome (DS)/Familial febrile seizures (FFS)/Generalized epilepsy with febrile seizures plus (GEFS+) [154,155] |
| GRIN2A | -0.41 | 5.2e-04 | -0.58 | 1.4e-08 | -0.29 | 6.0e-05 | Down | Focal epilepsy and speech disorder (FESD) [156,157] |
| MEF2C | -0.44 | 2.0e-04 | -0.66 | 1.3e-09 | -0.22 | 6.3e-04 | Down | Mental retardation [158] |
| FGF12 | -0.57 | 1.7e-06 | -0.85 | 2.7e-14 | -0.38 | 8.6e-07 | Down | EIEE [159,160] |
| GABRA1 | -0.68 | 2.5e-05 | -0.93 | 1.2e-11 | -0.23 | 1.5e-02 | Down | EIEE/CAE/Juvenile myoclonic epilepsy (JME) [161–163] |
| GABRG2 | -0.69 | 6.0e-06 | -0.82 | 5.1e-11 | -0.53 | 1.8e-08 | Down | FFS/GEFS+/CAE [64,164] |

\* FC is the Fold Change in Log2 scale.

### Intersection between SPD cluster in AD and epilepsy-related pathways

We performed multiple biological pathway analyses and identified epilepsy-related pathways, including Glutamatergic synapse, Dopaminergic synapse, and GABAergic synapse (Fig 3a), all involved in various neurological and psychiatric disorders [44–49]. These pathways include genes encoding glutamate and GABA receptors (*GRIN2A, SLC1A2, PLCB1, GRIN2B, SCN1A, GABRB3, GABRA1, and GABRG2*) (**Fig 3b**).

**Fig. 3:**
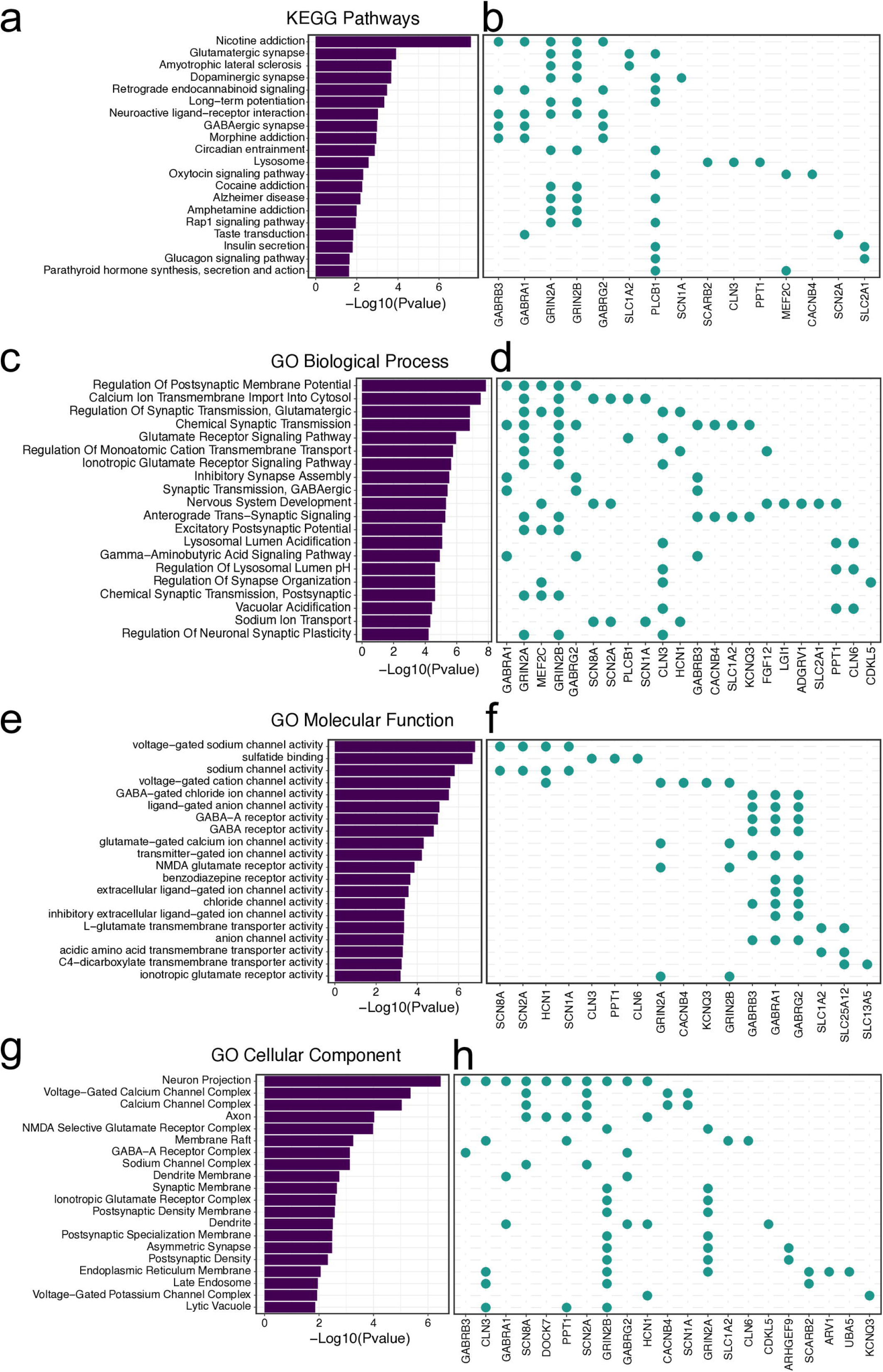
Intersection between SPD profile in AD and epilepsy-related pathways. (**a**) Barplot showing the top 20 KEGG pathways. (**b**) Circle plot showing the epilepsy-causing genes invovled in each pathway. (c,d,e,f,g, and h) Same as “**a**” and “**b**” but for GO biological process, GO molecular function, and GO cellular component respectively.

Additionally, we identified the Nicotine addiction pathway, which includes the genes *GABRB3*, *GABRA1*, *GRIN2A*, *GRIN2B*, and *GABRG2 (**Fig 3b**)*. Growing evidence shows that patients with epilepsy smoke at high, which may lead to an increase in seizure frequency [50–52]. Another significant pathway is circadian entrainment due to two glutamate ionotropic receptors (*GRIN2A*, *GRIN2B (***Fig 3b**). Increasing evidence shows the interaction between epilepsy and circadian rhythms [53].

Next, we looked at the Gene Ontology (GO) biological process pathways and observed that many were related to synaptic transmission, regulation, and glutamate signaling pathways (**Fig 3c**). These pathways were enriched in synaptic genes (**Fig 3d**). The sodium ion transport pathway is another significant one (**Fig 3c**). Many genetic mutations within genes that encode sodium channels have been identified in patients with epilepsy [54]. One of the top GO molecular function pathways identified were channel activity pathways (**Fig 3e**), including voltage-gated sodium channel activity, GABA-gated chloride ion channel activity, ligand-gated anion channel activity, glutamate-gated calcium ion channel activity, chloride channel activity, which are implicated in the pathogenesis of epilepsy [54–58]. The top GO cellular component pathways were related to channel/receptor complex and synaptic membrane-related pathways (**Fig 3g-h**). Altogether, these results show that the SPD cluster of AD is significantly enriched in epilepsy-related pathways.

### Epilepsy-causing gene dysregulation at multiple stages of AD

We selected the epilepsy-associated genes dysregulated in the SPD cluster and examined the effect size differences between the early-AD vs. control and the late-AD vs. control among the ROSMAP cohort (**Fig 4a-b**). Interestingly, we observed a concordant change of epilepsy-causing gene dysregulation in early and late AD compared to controls; however, the effect was more pronounced at later stages (**Fig 4b**, R^2^_adj_ = 0.94, p = 1.1×10^-15^, slope=1.2). This was consistent among 25 epilepsy genes including 23 down-regulated genes (*ARHGEF9*, *CACNB4*, *CDKL5*, *FGF12*, *GABRA1*, *GABRB3*, *GABRG2*, *GRIN2A*, *GUF1*, *HCN1*, *KCNQ3*, *MEF2C*, *NECAP1*, *NHLRC1*, *PPT1*, *PRICKLE1*, *SCARB2*, *SCN1A*, *SCN2A*, *SCN8A*, *ST3GAL5*, *STRADA*, *UBA5*) and two up-regulated genes (*CERS1* and *WWOX*) (**Fig 4b**). *WWOX* is associated with epileptic encephalopathy (EE) [59–63] (aka WOREE syndrome or WWOX-related epileptic encephalopathy), a severe type of epilepsy associated with severe cognitive and behavioral impairment starting in infancy and early childhood. Similarly, homozygous mutations in the *CERS1* gene have been linked to progressive myoclonic epilepsies Type 8 [41,42]. Three genes, *GABRG2*, *HCN1*, and *PRICKLE1*, had the most robust effect size (< -0.3 in both stages) among down-regulated genes (**Fig 4b**). Mutations in the *GABRG2* gene have been linked to multiple epilepsy syndromes, including simple febrile seizures (FS), childhood absence epilepsy (CAE), generalized epilepsy with febrile seizures plus (GEFS+), and Dravet syndrome (DS) [64]. Previous studies have shown that mutations in HCN1 can cause epilepsy through multiple HCN1 channel mechanisms, including loss-of-function (LOF) and gain-of-function (GOF) [65–67]. Similarly, previous work has identified mutations in the *PRICKLE1* gene associated with autosomal recessive progressive myoclonus epilepsy-ataxia syndrome [68,69].

**Fig. 4:**
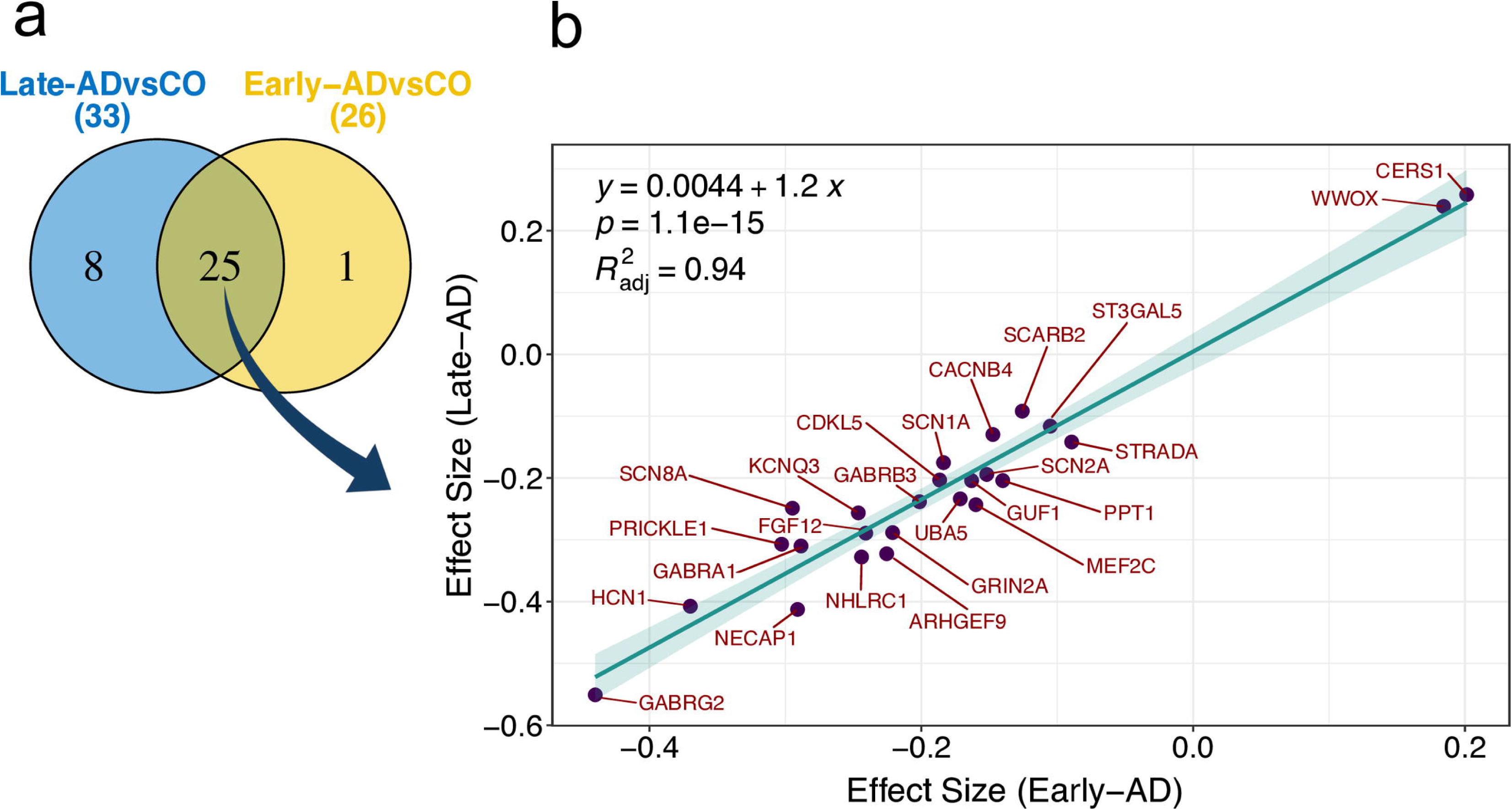
Epilepsy-causing gene are dysregulated at multiple stages of AD. **(a)** Venn diagrams showing the number of SPD dysregulated epilepsy-causing genes between early-AD vs control and late-AD vs control in the ROSMAP cohort, as well as the number of the common ones among the two comparisons. (b) Scatterplot showing the correlation of effect size between early-AD and late-AD for common SPD epilepsy-causing genes in the ROSMAP cohort.

## Discussion

The present study analyzed high-throughput transcriptomic data from sporadic AD cases and other AD cases with more severe cognitive profiles (SPD profile) across multiple AD cohorts and brain regions. We aimed to examine the relationship between molecularly defined profiles of AD and epilepsy-causing genes. Our findings corroborate the epilepsy-AD connection. In addition, we observed that these epilepsy-causing genes are mainly downregulated in AD earlier stages and correlate with AD progression. These results may indicate that epilepsy-causing genes are associated with both the onset and progression of AD, which could suggest that epileptic mechanisms might be present before or concurrently with the initial stages of AD. However, it remains uncertain which disorder precedes the other.

We found several epilepsy-causing genes/pathways dysregulated in the AD SPD profile, 16 of which were shared across the three cohorts of AD. Interestingly, 15 were consistently downregulated across the three cohorts, indicating that these genes probably encode proteins that present fundamental roles in cognitive function. We observed nine synaptic genes (*ARHGEF9*, *GABRA1*, *GABRB3*, *GABRG2*, *GRIN2A*, *HCN1*, *NECAP1*, *SCN1A*, *SCN8A*) highly expressed in iPSC-derived neurons, which may indicate synaptic dysfunction for both AD and epilepsy. *ARHGEF9* is a post-synaptic gene known to play a role in GABAergic synapses [70], and exhibits significant clinical heterogeneity, with epilepsy being a typical clinical phenotype [71–73]. *GABRA1*, *GABRB3*, and *GABRG2* are genes that encode GABAA receptor subunits. Disrupted GABAergic neurotransmission has been found in AD [74], and impaired GABAergic transmission might lead to the development of seizures in the early stages of AD through hippocampal hyperexcitability and an imbalance between excitation and inhibition [75]. We also identified *GRIN2A*, a gene encoding for the GluN2A subunit of the NMDA receptors [76]. Mutations in *GRIN2A* have been reported to be associated with multiple neurodevelopmental disorders, including epilepsy-aphasia spectrum (EAS), developmental delay (DD), and intellectual disability (ID) [76–78]. *GRIN2A* is downregulated in AD samples [79]. These results suggest that *GRIN2A* may have protective roles, and its downregulation could contribute to the AD-epilepsy connection.

The decreased expression of *HCN1* was shown to be associated with epilepsy [80], visual function [81,82] and learning and memory [83,84]. *NECAP1* isa gene that encodes for a protein involved in clathrin-mediated endocytosis [85], playing a fundamental role in the early stages of this process. It is critical for synaptic vesicle recycling, a key process in neurotransmission [86,87]. The loss of function of *NECAP1* results in severe infantile epileptic encephalopathy [85,88–90]. *SCN1A* and *SCN8A* encode two of the four voltage-gated sodium channels (VGSCs) primarily expressed in the central nervous system. Beyond their well-known association with epilepsy [91,92], *SCN1A* and *SCN8A* have also been frequently associated with ID and cognitive impairment [93–96], even in the absence of seizures [97,98]. Clinical screening of a family with Dravet Syndrome (DS) recently found an *SCN1A* missense mutation [99]. Years later, one family member, initially diagnosed with generalized epilepsy without ID, was diagnosed with AD, and a neuropathological examination confirmed the AD diagnosis by identifying NFT and Aβ pathology [99]. These findings align with our findings and suggest the connection between AD and DS patients, which often face early mortality and cognitive deficits.

We also identified *KCNQ3*, which encodes for the voltage-gated potassium channel subfamily Q member 3. Mutations in *KCNQ3* are associated with various forms of epilepsy, including benign familial neonatal epilepsy, infantile epilepsy, and epileptic encephalopathy with ID and cortical visual impairment [100–103]. Moreover, we identified *UBA5*, *PPT1*, *SLC25A12*, *MEF2C*, and *FGF12*, all of which have been reported to play a role in epilepsy and AD [104–125]. Notably, we found downregulation of *MEF2C*, which maintains proper physiological processes in multiple brain cell types [117].

Interestingly, *CERS1* was the only gene upregulated in the SPD cluster, which encodes Ceramide Synthase 1, an enzyme involved in ceramide biosynthesis [126]. It catalyzes the synthesis of C18 ceramide and is expressed in many tissues, predominantly in the CNS [126,127]. *CERS1* deficiency causes Progressive Myoclonic Epilepsy [41,42]. Increased cerebrospinal fluid levels and plasma C18 ceramide are associated with elevated AD markers, worse disease progression, and inflammation [128–130]. The decreasing plasma ceramide C18 concentration is also associated with improved cognitive performance [131]. Moreover, chronic insulin resistance, another feature of AD [132], increases ceramide production, further impairing insulin signaling and creating a vicious cycle with significant implications for AD [133]. *CERS1* is regulated by the PI3K/AKT signaling pathway [134], which is dysregulatedin AD [135]. Interestingly, disturbed insulin signaling, particularly the PI3K/AKT pathway, has also been proposed in epilepsy [19,136,137]. Hence, alterations in the regulation of *CERS1* and its function in producing ceramide emphasize the potential importance of ceramide metabolism in both epilepsy and AD.

We found larger effect sizes in epilepsy-causing genes at later AD stages which indicate that AD patients are at an increased risk for seizures in advanced disease stages [138]. Corroborating our findings, a recent study reported that subclinical epileptiform activity (SEA) increased in the AD continuum compared to controls, with its presence correlating with AD progression [139]. In these AD cases, SEA was associated with more severe visuospatial and attention impairments [139].

In a recent study, ∼90% of proteins altered in the hippocampus of epilepsy patients were also significantly changed in advanced AD [20]. Most of synapse and mitochondrial proteins were altered in the same direction in both conditions; many ribosomal proteins were changed in opposite directions [20]. Interestingly, some proteins reported by the authors overlap with the findings in our study, including *SCN2A* decreased in epilepsy and AD [20]. *SLC25A22*, another SLC25 mitochondrial carrier family member, was reduced in both disorders [20], aligning with the downregulation of *SLC25A12* observed in our study. *SCN2A*, one of the four VGSCs primarily expressed in the CNS, was decreased in their research [20], along with the downregulation of *SCN1A* and *SCN8A* observed here. These findings highlight common molecular pathways and potential therapeutic targets shared between epilepsy and AD, particularly in synaptic and mitochondrial function.

Although dysregulated epilepsy-causing genes might be unleashing AD pathogenesis, AD pathology may be triggering seizures along with disease onset. Thus, the dysregulation of epilepsy-associated genes could occur after AD pathology initiates. These two hypotheses underscore the complexity of the relationship between epilepsy and AD. A third and more plausible hypothesis is that the interrelation between the two suggests that either condition could precede the other, and the onset may vary between individuals.

## Conclusion

The intricate interaction between epilepsy and AD reveals a two-way relationship influenced by common genetic pathways and overlapping pathophysiological mechanisms. Our study shows that epilepsy-causing genes are disrupted in AD, particularly in later stages, indicating a potential role in the progression of the disease. These findings highlight the possibility that mechanisms involved in epilepsy may precede or co-occur with early AD pathology. At the same time, changes associated with AD may also worsen neuronal hyperexcitability and lead to seizures. The reduced transcript levels of synaptic genes such as *ARHGEF9*, *GABRA1*, *GABRB3*, and others indicate their crucial roles in cognitive function and potential association with epilepsy and AD. These findings support previous research linking disturbed GABAergic and glutamatergic neurotransmission to AD development and suggest that changes in these pathways contribute to the observed decline in cognitive function. Additionally, identifying down-regulated genes such as *SCN1A*, *SCN8A*, and *HCN1* emphasizes their dual roles in controlling neuronal excitability and synaptic function, implicating them in the progression of epilepsy and AD. Our results highlight the importance of genetic findings in the clinical context and indicate that AD patients with epilepsy tend to experience more severe cognitive impairments and faster disease progression. This supports clinical observations of increased seizure susceptibility in different stages of AD, particularly in advanced phases. It underscores the importance of early detection and management of epilepsy in AD patients. In addition, the shared genetic pathways between epilepsy and AD suggest potential therapeutic targets that could benefit both conditions. This highlights the necessity for integrated treatment strategies. Our findings contribute to a deeper understanding of the connection between epilepsy and AD, although the exact order in which they overlap remains unclear. We believe that the development of epilepsy and AD may vary among individuals, underscoring the importance of personalized approaches, particularly leveraging genetic technology. The comprehension of these mechanisms will facilitate the development of targeted treatments addressing shared genetic vulnerabilities, thereby potentially improving outcomes for individuals affected by both epilepsy and AD. The results of the Levetiracetam for Alzheimer’s Disease-Associated Network Hyperexcitability (LEV-AD) trial provide a hint that stratification based on the epileptiform phenotype in AD could be crucial, particularly for developing targeted therapies to reduce network hyperexcitability.

## Data Statement

Transcriptomics raw data from MSBB study are publicly available at Synapse under Synapse IDs syn3157743 (https://www.synapse.org/#!Synapse:syn3157743). Transcriptomics raw data from ROSMAP study are publicly available at Synapse under Synapse IDs syn17008934 (https://www.synapse.org/#!Synapse:syn17008934).

## CRediT authorship contribution statement

AEM data curation, formal analysis, methodology, software, original draft writing, review & editing. SB participated in funding acquisition. SB, MS, and DP resources and conceptualization. SSA and NGC participated in writing, reviewing & editing the first and last versions of the manuscript. BAB participated in conceptualization, funding acquisition, methodology, project administration, supervision, writing – review & editing of the first and last versions of the manuscript.

## Acknowledgment

We thank all the participants, their families, the many involved institutions, and their staff, whose help and participation made this work possible.

